# Association Between Sampling Method and Covid-19 Test Positivity Among Undergraduate Students: Testing Friendship Paradox in Covid-19 Network of Transmission

**DOI:** 10.1101/2020.12.14.20248144

**Authors:** Sina Kianersi, Yong-Yeol Ahn, Molly Rosenberg

## Abstract

In November 2020, we conducted a cross sectional study to implement and test the method of acquaintance sampling (randomly sampling friends of randomly sampled individuals) in detecting students with higher probability of COVID-19 positivity. Overall, 879 students were randomly sampled and participated in this study. In an online survey, the randomly sampled participants nominated a friend, and reported their own and their nominated friend’s COVID-19 status. Nominated friends were about 1.64 (95% CI: 1.33, 2.00) times more likely to have ever been infected with COVID-19, compared to randomly sampled students. Our study corroborates the effectiveness of acquaintance sampling for identifying members of networks with higher COVID-19 risk. These findings could be useful for university policy makers when developing mitigation testing programs and intervention strategies against COVID-19 spread.

## Introduction

Coronavirus Disease 2019 (COVID-19) has disproportionately affected college student populations (1). Because of the important roles of colleges in education, regional economy, and safety of surrounding areas, the development of effective interventions and prevention programs to reduce COVID-19 transmission among college students is crucial. Identifying students who are more likely to get COVID-19 could improve the disease containment efforts, such as intervention programs, surveillance testing, and even vaccination strategies, and make them more efficient and effective. “Friendship Paradox” (2), a fundamental concept in network science, could potentially help to identify students with higher COVID-19 positivity risk.

Studies on epidemic spreading in networks have established two fundamental mathematical principles. First, on average, the neighbors (or friends) of a randomly selected node have higher degree than the node (“Friendship Paradox”), where the nodes are people and the degree is the number of acquaintances (2). Secondly, following this first principle, people (nodes) with higher degree are preferentially attacked in an epidemic (3). These principles underpin the methodology of acquaintance sampling, where randomly sampled individuals are asked to nominate friends. The friends, should, on average, have higher disease risk than the randomly sampled population.

Acquaintance sampling has been used to sample the high-degree nodes in general populations (4, 5) and consists of two steps: Step 1) select a random sample of population, Step 2) randomly select a link (in our case a friend) of the randomly sampled individual in step 1. This method does not require any knowledge of the underlying contact network structure and can be in principle implemented for any large general population (4). Previously, this method has successfully been tested for early detection of flu outbreaks (6). However, it is not yet established whether acquaintance sampling can be successfully applied to COVID-19 pandemic, particularly under a highly disrupted physical contact network structure due to other intervention measures.

The aim of the current study was to implement and test the acquaintance sampling technique in detecting students with higher probability of COVID-19 positivity. We hypothesized that students sampled in step 2 (i.e., nominated friends) would be more likely to have COVID-19 compared to students sampled in step 1 (i.e., a random sample of students).

## Methods

From September to November 2020, we conducted a randomized controlled trial (RCT) to test whether receiving serological COVID-19 test results influenced COVID-19 protective behaviors. Details about the study methods are included in other study reports (7). In the current study, we report our findings about the cross-sectional data that we collected in the endline survey of the RCT. The Indiana University Office of Research Compliance approved the study protocol (protocol #2008293852).

Overall, we randomly sampled 4,069 Indiana University (IU) undergraduate students and invited them to participate in the study. The endline survey was an online survey on REDCap and included questions about student’s previous positive testing history (if the student has ever been tested positive for COVID-19). We also asked students to nominate one of their friends (step 2 of the sampling method) using the following question;

- Please think of one of your friends in IU whom you talked to recently; Please type your friend’s initials in the box.

Next, we asked students to report if the friend that they had selected in the previous question had ever tested positive for COVID-19.

- Has your friend with the initials “[initials entered for previous question]” ever been infected with COVID-19?

We used Poisson regression models with a robust error variance (8) to assess the crude association between sampling method (randomly sampled vs. nominated friend) and the self-reported COVID-19 test results (positive vs. negative).

## Results

Overall, 879 randomly sampled students self-reported their COVID-19 test results, and 843 students nominated a friend and reported their nominated friends’ COVID-19 status. In the random sample, the prevalence of COVID-19 positivity was 14%. This was 23% among the nominated friends. Nominated friends were 1.64 (95% CI: 1.33, 2.00) times more likely to have ever been infected with COVID-19 (Table 1).

**Table 1.** Association between sampling method and COVID-19 self-reported test results

|  |  |  |  |  |
| --- | --- | --- | --- | --- |
|  | Total<br>N= 1,722 | Random sample<br>N=879 | Nominated<br>Friend<br>N=843 | Prevalence Ratio<br>and 95 % CI |
| COVID-19 Self-Reported Result |  |  |  |  |
| Positive | 316 (18.4%) | 123 (14.0%) | 193 (23.0%) | **1.64 (1.33, 2.01)** |
| Negative | 1406 (81.7%) | 756 (86.0%) | 650 (77.1%) | Ref. |

## Discussion

We found a positive association between the sampling method and COVID-19 positivity. Nominated friends were more likely to have had COVID-19 compared to randomly sampled students.

The concept of the friendship paradox and the related sampling technique have been previously used for early detection of seasonal flu epidemics among university students (6). Moreover, it has been shown that this sampling technique could be used for targeted immunization against epidemic attacks (9). We found that there is a strong association between sampling method and COVID-19 positivity among college students. Epidemiologists and policy makers might find this information useful when designing surveillance testing programs.

The primary limitation of the current study is that we could not contact the nominated friends to collect data about their actual COVID-19 testing history because this was an opportunistic study that leveraged a larger RCT. Participants might had imperfect and out of date information about their friends’ COVID-19 status. This particularly could have caused underestimation of the true COVID-19 prevalence among the nominated friends. Moreover, because students were participating in a study about COVID-19, it might have primed them to recall their friends who had been tested positive in the past. This selection bias might have caused an overestimation of the true prevalence of COVID-19 positivity among nominated friends.

Despite these limitations, our study bolsters the strength of the acquaintance sampling method in detecting those who are more likely to contract the disease early. These findings could be useful for university policy makers when developing mitigation testing programs and intervention strategies against COVID-19 spread. We suggest future studies assess the feasibility of this sampling method in mitigation/surveillance testing and immunization strategies at university level.

## Data Availability

Data are available upon request.

